# The risk of symptomatic reinfection during the second COVID-19 wave in individuals previously exposed to SARS-CoV-2

**DOI:** 10.1101/2021.04.14.21255502

**Authors:** Mattia Manica, Serena Pancheri, Piero Poletti, Giulia Giovanazzi, Giorgio Guzzetta, Filippo Trentini, Valentina Marziano, Marco Ajelli, Maria Grazia Zuccali, Pier Paolo Benetollo, Stefano Merler, Antonio Ferro

## Abstract

To what extent infection with SARS-CoV-2 protects against subsequent reinfection or symptomatic reinfection is still unclear. In this cohort study, we analyzed surveillance records of COVID-19 cases identified between June 2020 and January 2021 in five Italian municipalities, where 77.7% of the entire population was screened for IgG antibodies in May 2020. We compared the risk of observing symptomatic infections in two mutually exclusive groups defined by the initial serological response. We estimated that the cumulative incidence of identified symptomatic infections in the IgG negative and positive cohorts was 2.67% (95%CI: 2.12% – 3.37%) and 0.14% (95%CI: 0.04% – 0.58%), respectively. The adjusted odd ratio of developing symptomatic infection in individuals previously exposed to SARS-CoV-2 was estimated at 0.054 (95%CI: 0.009 - 0.169). Quantifying protective immunity against COVID-19 disease elicited by natural infection with SARS-CoV-2 is essential to inform strategies for controlling the pandemic in the forthcoming months.

## Main

Infection from SARS-CoV-2 is expected to provide temporary protective immunity against subsequent reinfection or against the risk of disease following reinfection episodes ^1,2^. Published evidence indicate that more than 90% of individuals develop IgG and neutralizing antibodies following primary infection, but that antibody titers may wane rapidly over time, particularly in mild and asymptomatic patients ^2,3^. Sporadic episodes of SARS-CoV-2 reinfection have been documented ^2,4–7^. However, to what extent and for how long natural infection provides protective immunity from SARS-CoV-2 is still debated.

Recent estimates suggest 80-85% protection from reinfection ^8,9^ and 99% against symptomatic disease ^10^ up to six months from the first infection. However, follow-up studies comparing infections in recovered individuals with well-matched naive individuals are still lacking ^2^. Cohort studies conducted so far mainly relied on the comparison of infection rates among individuals who had a previous PCR result. Due to the limited testing of asymptomatic and pauci-symptomatic, this approach may under ascertain individuals who have already experienced the infection in the past. Combining surveillance data with extensive serological screening applied to the general population could help reducing biases in the assessment of the risk of reinfection.

We analyzed five Italian municipalities within the Autonomous Province of Trento, Italy, where an IgG serological screening aimed at covering the entire adult resident population was conducted between May 5 and 15, 2020. These municipalities were selected as those showing the highest cumulative case incidence in the province during the first COVID-19 wave ^11^ (ranging between 18.7 and 27.6 per 1,000 individuals). Serological tests were performed using Abbott SARS-CoV-2 IgG chemiluminescent assays and analyzed on the Abbott Architect i2000SR automated analyzer (Abbott Diagnostics, Chicago, IL, USA) ^11^. The employed assay detects IgG directed against the SARS-CoV-2 nucleocapsid protein, measured as a Relative Light Unit (RLU), which is considered as a proxy of the concentration of IgG antibodies to SARS-CoV-2 in the sample. Serological results are provided as the ratio between sample RLU and the calibrator mean chemiluminescent signal from three calibrator replicates. Results are interpreted as positive when this ratio is ≥1.4 and negative when <1.4 ^11^. The serological screening involved 6,074 individuals (median age 50; IQR: 32-63), representing 77.1% of the resident population (Table 1). Of these, 1,402 (23.1%) resulted positive for IgG antibodies. Among IgG positive individuals, 12.8% (i.e. 180 infections) had a confirmed naso-oropharyngeal swabs test before the serological screening, a percentage consistent with the ascertainment rate estimated for Italy during the first COVID-19 epidemic wave ^12^. Between June 1, 2020 and January 31, 2021, regular surveillance activities conducted to control COVID-19 in the province identified 221 new positive SARS-CoV-2 infections among study participants. Positive cases were ascertained by using either RealTime SARS-CoV-2 assay on naso-oropharyngeal swabs (detectability per ml of UTM buffer 250 copies) or rapid antigenic test (sensitivity >90%, specificity >97%). Out of 221 confirmed cases, 124 were symptomatic (Table 1). Symptomatic infections were defined as positive participants having fever and either cough or at least two of the following symptoms: widespread myalgia, headache, dyspnoea, pharyngodynia, diarrhea, nausea/vomiting, anosmia/ageusia, asthenia.

**Table 1.**
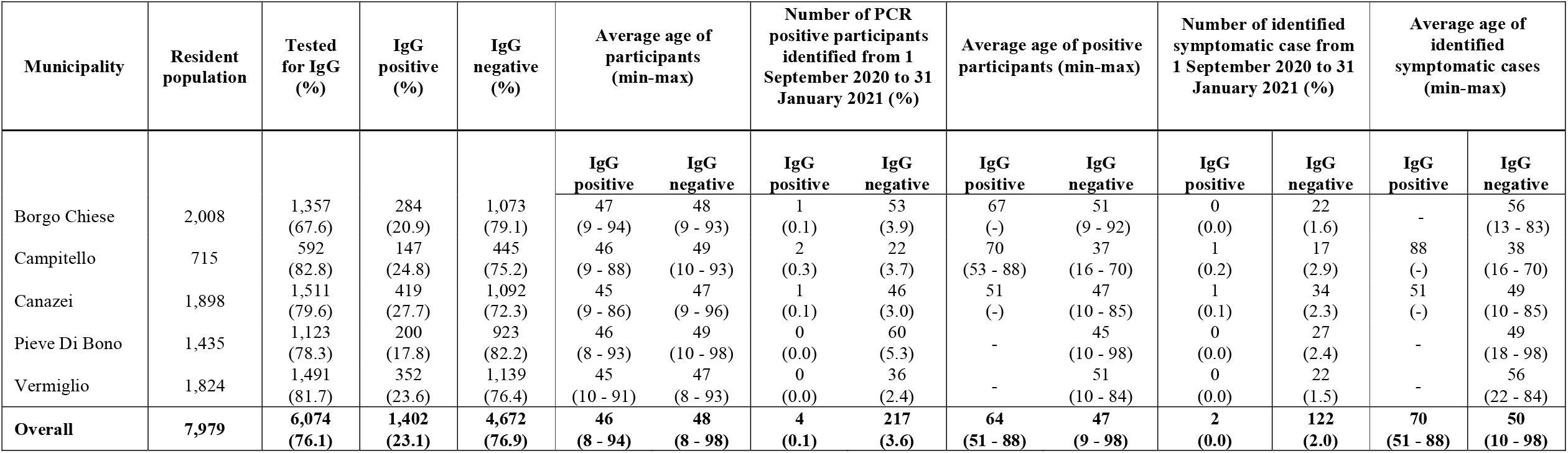
Descriptive statistics.

Four cases were identified among participants who tested positive to IgG in May 2020; two of them were symptomatic. Both these cases were males ascertained in December 2020, who requested to be tested after symptoms onset. The older patient (88 years) was admitted to a hospital but did not require mechanical ventilation or admission to an intensive care unit. The younger patient (52 years) was a mild case who was isolated and treated at home.

We estimated the relative risk of developing a symptomatic infection for participants who tested positive for IgG antibodies in May 2020 compared to those who resulted IgG negative to SARS-CoV-2 infection. To do this, we applied a generalized linear mixed model with logit link, defining the dependent variable as the confirmation of a symptomatic infection occurred between June 1, 2020 and January 31, 2021 and using the participant age and IgG binary result obtained in May 2020 (positive vs negative) as independent variables. Age was standardized by subtracting the mean and dividing by standard deviation. The municipality of residence was considered as a random effect to account for possible heterogeneity in exposure to SARS-CoV-2 across different geographical areas. The cumulative incidence of identified symptomatic infections over the observation period was 2.67% (95%CI: 2.12% – 3.37%) in the IgG negative group and 0.14% (95%CI: 0.04% – 0.58%) in the IgG positive group. The adjusted odds ratio of being confirmed as a symptomatic SARS-CoV-2 infection in IgG positive relative to IgG negative participants was 0.054 (95%CI: 0.009-0.169, see Table 2).

**Table 2.** Effect sizes estimated by applying a generalized linear mixed model to the likelihood of observing SARS-CoV-2 symptomatic infections in individuals previously tested for IgG antibodies.

| Variable | Estimate | std. error | Z value | p-value |
| --- | --- | --- | --- | --- |
| (Intercept) | -3.62 | 0.118 | -30.67 | <0.00001 |
| IgG Positive | -2.93 | 0.713 | -4.11 | 0.00004 |
| Standardized age | 0.11 | 0.091 | 1.23 | 0.21984 |
| Random effect SD | 0.16 |  |  |  |

The conducted analysis confirms the hypothesis that the likelihood of experiencing SARS-CoV-2 symptomatic infection is greatly reduced in individuals already infected in the previous 8-10 months^11^. In line with what observed elsewhere ^7–9,13,14^, our findings suggest that the relative risk of symptomatic infection for individuals who previously tested positive to IgG antibodies compared to seronegative subjects is less than 6%.

Our results should be interpreted in light of the following limitations. First, as study participants were not enrolled for regular PCR testing following the IgG survey, a fraction of symptomatic reinfection events occurred in the population might have been missed by syndromic surveillance. Second, the observed reinfection events depend not only on the duration and amount of protection against reinfection, but also from the individual number of contacts and temporal changes in the prevalence of infection in the general population. Given the small size of the municipalities considered in our study, we assumed that exposure to SARS-CoV-2 was similar for individuals of the same age and residing in the same municipality. Third, the analyzed data do not provide any information about the potential presence of SARS-CoV-2 lineages emerged in recent months. Therefore, estimates obtained here may not apply to SARS-CoV-2 variants that are quickly replacing historical lineages circulating in 2020 ^2^. Finally, the study design was not suitable to disentangle whether the infection from historical lineages of SARS-CoV-2 would either protect individuals against reinfection (sterilizing immunity) or from symptoms development during the reinfection episode, as a large share of asymptomatic infections^15^ could have remained undetected by the surveillance system (87% before May 2020^10^).

The major strength of the proposed analysis is that study participants cover 78% of residents of 5 municipalities, providing a comprehensive view of infection risks in the general population. In addition, individuals who were previously exposed to SARS-CoV-2 were identified via IgG serological testing, therefore reducing biases caused by under ascertainment of infection episodes in asymptomatic and mild disease cases. Further studies are still needed to quantify sterilizing immunity against SARS-CoV-2 and its duration, to explore whether immune responses mounted following initial infection can prevent possible onwards transmission, and to investigate cross-protection across different SARS-Cov-2 lineages.

## Data Availability

The data used for the analysis will be made openly available both upon acceptance of the manuscript.

## Ethical statement

Informed consensus for blood collection was obtained from all the participants. The study was approved by the Ethics Committee of the Istituto Superiore di Sanità (Prot. PRE BIO CE n.15997, 04.05.2020).

## Author contributions

PP,SM,AF conceived and designed the study. MM performed the analysis. SP,GGi,MGZ,PPB,AF collected data. MM,PP wrote the first draft. All authors contributed to data interpretation, critical revision of the manuscript and approved the final version of the manuscript.

## References

1. Stokel-Walker, C. What we know about covid-19 reinfection so far. BMJ n99 (2021) doi:10.1136/bmj.n99.

2. European Centre for Disease Prevention and Control. Risk of SARS-CoV-2 transmission from newly-infected individuals with documented previous infection or vaccination. (ECDC, 2021).

3. Chia, W. N. et al. Dynamics of SARS-CoV-2 neutralising antibody responses and duration of immunity: a longitudinal study. Lancet Microbe S2666524721000252 (2021) doi:10.1016/S2666-5247(21)00025-2.

4. Adrielle dos Santos, L. et al. Recurrent COVID-19 including evidence of reinfection and enhanced severity in thirty Brazilian healthcare workers. J. Infect. 82, 399–406 (2021).

5. Elzein, F. et al. Reinfection, recurrence, or delayed presentation of COVID-19? Case series and review of the literature. J. Infect. Public Health S187603412100006X (2021) doi:10.1016/j.jiph.2021.01.002.

6. Iwasaki, A. What reinfections mean for COVID-19. Lancet Infect. Dis. 21, 3–5 (2021).

7. Zhang, J. et al. COVID-19 reinfection in the presence of neutralizing antibodies. Natl. Sci. Rev. nwab006 (2021) doi:10.1093/nsr/nwab006.

8. Hansen, C. H., Michlmayr, D., Gubbels, S. M., Mølbak, K. & Ethelberg, S. Assessment of protection against reinfection with SARS-CoV-2 among 4 million PCR-tested individuals in Denmark in 2020: a population-level observational study. The Lancet S0140673621005754 (2021) doi:10.1016/S0140-6736(21)00575-4.

9. Hall, V. J. et al. SARS-CoV-2 infection rates of antibody-positive compared with antibody-negative health-care workers in England: a large, multicentre, prospective cohort study (SIREN). The Lancet S0140673621006759 (2021) doi:10.1016/S0140-6736(21)00675-9.

10. Abu Raddad, L. J. et al. Assessment of the risk of SARS-CoV-2 reinfection in an intense reexposure setting. http://medrxiv.org/lookup/doi/10.1101/2020.08.24.20179457 (2020) xdoi:10.1101/2020.08.24.20179457.

11. Stefanelli, P. et al. Prevalence of SARS-CoV-2 IgG antibodies in an area of northeastern Italy with a high incidence of COVID-19 cases: a population-based study. Clin. Microbiol. Infect. S1198743X20307096 (2020) doi:10.1016/j.cmi.2020.11.013.

12. Marziano, V. et al. Retrospective analysis of the Italian exit strategy from COVID-19 lockdown. Proc. Natl. Acad. Sci. 118, e2019617118 (2021).

13. Harvey, R. A. et al. Association of SARS-CoV-2 Seropositive Antibody Test With Risk of Future Infection. JAMA Intern. Med. (2021) doi:10.1001/jamainternmed.2021.0366.

14. Lumley, S. F. et al. Antibody Status and Incidence of SARS-CoV-2 Infection in Health Care Workers. N. Engl. J. Med. 384, 533–540 (2021).

15. Poletti, P. et al. Association of Age With Likelihood of Developing Symptoms and Critical Disease Among Close Contacts Exposed to Patients With Confirmed SARS-CoV-2 Infection in Italy. JAMA Netw. Open 4, e211085 (2021).

